# Exploring the changing association between parental and adolescent fruit and vegetable intakes, from age 10 to 30 years

**DOI:** 10.1101/2024.01.26.24301777

**Authors:** Tanya Braune, Prof Jean Adams, Dr Eleanor M. Winpenny

## Abstract

**Background:** Adolescence is a pivotal developmental stage, where escalating rates of overweight and obesity have raised concerns about diet quality and its association with adverse health outcomes. Parents are known to have considerable influence on childhood diet, but how this influence changes as adolescents mature is unknown. This study investigates the association between parental fruit and vegetable (FV) intake and adolescent FV consumption, exploring how this changes across adolescence and when children leave home.

**Methods:** Adolescents aged 10-30 years (n=12,805) from the UK Household Longitudinal Study (UKHLS), and their parents, reported FV intakes every 2 years. Multilevel linear regression models were fitted to assess associations between parental and adolescent FV intakes, investigating interactions with age and living arrangement, and adjusting for sociodemographic covariates.

**Results:** Parental FV intake was positively associated with adolescent FV intake (β=0.20 [95%CI:0.19,0.22] portions/day), with the strength of this association lowest during early adolescence (10-14 years) and peaking at 17-18 years (β=0.30 [95%CI: 0.27,0.33] portions/day). When adolescents no longer lived in the parental home, the association of parental FV intake with their own FV consumption decreased, but a positive association was maintained up to age 30 years.

**Conclusions:** Our findings emphasise the enduring effect of parental modelling on dietary choices, highlighting the potential for interventions to promote increased FV intake, acknowledging the lasting influence of parental diet, even beyond the confines of the parental home.

## 1. Introduction

Adolescence is a significant developmental life stage and adequate nutrition during this time is key to future health outcomes and establishing dietary habits for adulthood^1,2^. There are ongoing concerns around the rapidly increasing prevalence of overweight and obesity, particularly among children and adolescents^3–5^. Poorer diet quality is associated with higher fat mass^6,7^, adiposity^8^, elevated body mass index (BMI)^9^ and cardiometabolic risk factors^10^ in children and adolescents, which have been shown to persist into adulthood, posing a continued risk for chronic disease^11–18^. For example, in coronary artery disease, the development of atherosclerotic plaques associated with the disease are seen to be initiated during adolescence and young adulthood, starting at age 15^19^. With growing evidence linking diet with overweight and obesity and chronic disease^20^, it is of particular importance to understand how diets are established in adolescence and the key influences involved.

Higher intake of fruit and vegetables (FV) is a major component of a healthier diet, with worldwide dietary recommendations and the World Health Organization (WHO) encouraging a minimum intake of 400g or five portions per day^21,22^ due to the inverse relationship seen between FV intake and obesity^23–25^ as well as chronic disease^11–14,17^. Moreover, increasing FV consumption has been shown to help with weight loss among people with overweight and obesity^26^. The National Diet and Nutrition Survey (NDNS) has regularly reported that FV consumption in the United Kingdom (UK) is below the recommended daily intake of at least five portions^12,17^, which was shown to be lowest in children aged 11-18 years at 2.9 portions per day in the 2016-2019 NDNS report^27^ and remaining low at 2.8 portions per day in 2020^28^. This was further highlighted by results from Health Survey England, showing that young people aged 16-24 years have consistently consumed less FV than older adults^29^. Public health efforts in the UK such as the launch of the five-a-day campaign and the Eatwell plate, have failed to significantly increase average intake to the recommended level, particularly in young people^29^.

Adolescents and young adults experience a high number of changes in their social and physical environments, particularly during the transition from adolescence to early adulthood^30^. Diet in adolescents has been shown to be influenced by the behaviours of peers and parents^31–33^. Parents are responsible for the majority of their child’s food choices throughout their early lives and food preferences in childhood are often established by what has been modelled and provided by their caregivers^34^. As children enter adolescence, they begin to gain higher control over their diet with less supervision^34^. However, research more often concentrates on younger children and parenting practices during earlier childhood^30^ and there is less understanding of how parents influence their children’s diets in adolescence and early adulthood. Existing literature has shown an overall positive association between parental and adolescent intake when considering healthier (e.g. FV) or less healthy (e.g. sugar-sweetened beverages or snacking) measures^31,35–39^. However, the current literature is limited mostly to cross-sectional analyses, with only four longitudinal analyses^38,40–42^ that focussed on earlier adolescence (10-15 years), which cannot capture the change over time of these influences during the transition from late adolescence into young adulthood^43^. Importantly, it is unclear how the influence of parents in adolescence is maintained into adulthood.

During the transition from adolescence to early adulthood, individuals will often experience changes in their living arrangement. Despite some evidence that behaviours introduced during adolescence can be followed into adulthood^30^, there has been limited exploration into how a change in living arrangement influences dietary habits longer term. Adolescents’ and young adults’ diets change, depending on whether they live in the parental home or not^44–48^. For example, Winpenny et al. found that FV consumption frequency decreases on leaving the parental home^44^. This further suggests that parental influence changes, not only as adolescents age, but as they gain independence over their food choices when they do not live in the parental home. However, there is a lack of evidence quantifying how the association between parental and adolescent intake changes.

The key objectives of this study were to explore the association between parental FV intake and adolescent FV intake, how this association changes with adolescent age, and whether it is dependent on adolescents living in the parental home. This analysis used a large ongoing longitudinal panel study in a UK population. Sawyer et al. defined the period of adolescence as 10 to 24 years^49^. The Office for National Statistics (ONS) found that in 2017, by the age of 23 years over half of young people had left home in the UK^50^. For the purposes of this study, the age group of interest extends up to age 30 years and will henceforth be referred to as ‘adolescents’, to assess the association between parental FV intake and adolescent FV intake after the adolescents no longer live in the parental home, and explore how this association may persist over the longer term.

The objectives were achieved through answering the following research questions:

1. How is parental FV consumption cross-sectionally associated with adolescent FV consumption across adolescence and early adulthood (10-30 years)?
2. How does the association between parental and adolescent FV consumption differ across adolescent age?
3. How does the association between parental and adolescent FV consumption change when adolescents do not live in the parental home?

### 2. Methods

#### 3.1. Study design and population

Data for this analysis were drawn from the UK Household Longitudinal Study (UKHLS), a longitudinal household survey that has collected a wide range of data each year, since 2009, in approximately 40,000 UK households^51^. Household members aged 16 years and over completed the adult questionnaire and FV data was collected in waves 7 (2015-2016), 9 (2017-2019), 11 (2019-2021) and 13 (2021-2023). The youth questionnaire is completed by household members age 10-15 years and FV intake data was collected in waves 8 (2016-2017), 9 (2017-2019), 11 (2019-2021) and 13 (2021-2023). Each individual in the dataset is linked to others in their household and they continue to be invited to participate in the survey even if they leave the household. In this analysis, we include all adolescents between ages 10 and 30 years at any of the waves in which FV data was collected: adult (16-30 years) waves 7, 9, 11, 13 and youth (10-15 years) waves 8, 9, 11 and 13^51^. This resulted in an accelerated cohort design as depicted in Figure 1, which includes details of age of adolescent participants at each wave. Further details on the UKHLS sampling and survey methods can be found elsewhere^52^.

#### 3.2. Survey and Measures

##### Outcome: adolescent FV intake (portions per day)

In the adult questionnaire, fruit and vegetable intake frequency (days/week) and amount (portions/day) were self-reported separately (described in Table 1) at every odd wave starting at wave 7 (waves available: 7, 9, 11 and 13). A numerical average value was assigned to each frequency category as follows: never = 0 days, 1-3 days = 2 days, 4-6 days = 5 days, and every day = 7 days. This was then multiplied by the reported amount (in portions) to obtain portions per week of fruit and vegetables. Portions per day were calculated by dividing this value by seven and fruit and vegetables were combined into one variable by adding together the portions per day intake for each, to obtain the final intake variable of FV portions per day.

**Table 1:** Dietary survey measures used in UKHLS

|  | Measure | Question | Possible answers |
| --- | --- | --- | --- |
| Adult questionnaire | Fruit intake frequency | "Including tinned, frozen, dried and fresh fruit, on how many days in a usual week do you eat fruit?" | Never<br>1 - 3 Days<br>4 - 6 Days<br>Every day |
|  | Fruit amount | "On the days when you eat fruit, how many portions (e.g. an apple, an orange, some grapes) do you eat?" | Whole numbers |
|  | Vegetable intake frequency | "Including tinned, frozen and fresh vegetables, on how many days in a usual week do you eat vegetables? Do not include potatoes, crisps or chips." | Never<br>1 - 3 Days<br>4 - 6 Days<br>Every day |
|  | Vegetable amount | "On the days when you eat vegetables, how many portions (i.e. 3 heaped tablespoons) do you eat? Please do not include potatoes." | Whole numbers |
| Youth questionnaire | Fruit and vegetable daily intake | "How many portions of fresh fruit or vegetables do you eat on a typical day? One portion is one piece of fruit or one serving of a vegetable or salad item" | 5 or more portions<br>3-4 portions<br>1-2 portions<br>None |

In the youth questionnaire, participants self-reported their combined fruit and vegetables intake (see Table 1) at waves 8, 9, 11 and 13. The responses were reported in portions per day and therefore, a numerical average value was assigned to each category as follows: 5 or more portions = 5.5 portions, 3-4 portions = 3.5 portions, 1-2 portions = 1.5 potions, and None = 0, to obtain FV portions per day for analysis.

##### Exposure: parental FV intake (portions per day)

The main exposure was parental FV intake in portions per day. Adolescent data were matched to parental data using cross-wave identifiers provided by UKHLS, for their biological, step or adoptive parents, where available. Parental FV intake was calculated for each parent in the same way as adolescent FV intake using the adult questionnaire (outlined above). Then, an average parental value was calculated for daily FV intake. If one of the parents was missing, the intake variables were based only on data from one parent.

##### Confounders

A directed acyclic graph (DAG) was created to map likely confounders and the direction of the relationships between variables (see Figure 2). Based on this, the following confounders were adjusted for in the analyses: sex, ethnicity, parental education, gross household income and geographic region.

Ethnicity was self-reported by respondents in both adult and youth questionnaires or reported by household members or derived from ethnic group of their biological parents when missing, with priority given to self-reported information. There were 18 possible ethnicity categories, which were collapsed into two categories for analysis: 1) White and 2) non-White.

In the adult questionnaire, highest qualification achieved is collected through participant self-report and updated every year to include the most recent qualification. For this analysis, parental education was derived from the highest qualification between both parents. Of the six possible qualifications, a binary variable was created for parental education to include 1) degree level and above and 2) no degree (any other qualification below degree level, including no qualification).

Detailed information on monthly incomes were self-reported by every adult member of the household (aged 16 years and over) at each wave. For this analysis, the sum of the total net monthly household income without deductions was used and adjusted for household composition using an equivalence scale provided by UKHLS and reported in British Pound Stirling (£) per month.

Geographic region was self-reported and included 12 categories: North East, North West, Yorkshire and the Humber, East Midlands, West Midlands, East of England, London, South East, South West, Wales, Scotland and Northern Ireland.

##### Moderators

For the second and third research questions, we tested two potential moderators of the relationship between adolescent and parental FV intakes: 1) adolescent age and 2) whether or not the adolescent was living in the parental home. These were included as interactions with parental FV intake.

Age, in completed years, was derived based on reported date of birth and interview date. For the analysis, age was categorised in 2-year age bins (apart from age 10, to accommodate for the odd number of ages) in order to identify non-linear associations between parental intake and adolescent intake by age category.

Whether or not an adolescent was living in the parental home was already a binary variable, with possible responses being yes or no. This was determined by the UKHLS field team during the follow-up process. Adolescents were considered to no longer be living in the parental home if, at the time of the interview, they were temporarily absent (e.g. at college or university), in which case they would be considered a non-resident household member, or if they had left the household and were considered to be a split off household. Due to the household sampling method of UKHLS, any adolescent included in the analysis will have lived in their parental home at some stage.

##### Missing values and outliers

We selected participants aged 10-15 years in Waves 8, 9, 11 and 13 and aged 16-30 years in Waves 7, 9, 11 and 13 that had data available for their own FV intake, as well as for at least one of their biological, step or adoptive parents. The original sample of participants aged 10-15 years consisted of 5,784 respondents to the youth questionnaire and participants aged 16-30 years consisted of 13,532 respondents to the adult questionnaire (n=19,316). Of the original sample, 2,187 adolescents did not have FV intake data and were excluded from analysis. Of those that did have FV intake data available, 4,373 did not have parental FV intake data and therefore, these were not included in the final analytical sample. The key reason for missing FV intake values for both adolescents and parents was because they were not present for data collection and therefore, did not report their FV intake in that data collection wave. Additionally, not all participants in the age range had complete parental data for various reasons (e.g. parents were not part of the original sample). For covariates, there were 326 missing values for parental education after backfilling any missing values from previous waves (1-5). There were also 25 missing values for ethnicity, 4 for living in the parental home, 531 for net household income, and 12 for geographic region. Given the low proportion of individuals with missing covariate data in comparison to the sample size, these individuals were excluded from the analysis.

For FV intake data, outliers were assessed and data removed for values that were greater than three times the interquartile range of intake. A total of 593 data points for adolescent and parental intake of FVs (in youth and adult questionnaires) were excluded using this method. After data cleaning, the final analytical sample included 12,805 adolescent participants (66% of the original sample) consisting of 26,687 observations across four waves of data.

#### 3.3. Statistical Analysis

R statistical software^53^ (Version 2023.06.2+561) was the primary software used for statistical analysis. Python 3^54^ was used for data pre-processing. Descriptive statistics were computed for covariates, using frequency/percentage and mean/standard deviation where appropriate (see Table 2).

**Table 2:** Descriptive statistics of adolescent demographics (n = 12,805 adolescents, n = 26,687 observation

| | n (%) / mean ± SD |
| --- | --- |
| Age | 17.4 ± 5.6 |
| **Sex** |  |
| Female | 6,604 (52) |
| Male | 6,201 (48) |
| **Ethnicity** |  |
| White | 9,185 (72) |
| Non-white | 3,620 (28) |
| **Parental highest qualification** |  |
| Degree | 6,638 (52) |
| No degree | 6,167 (48) |
| **Income** |  |
| Net adjusted household income (£ per month) | 1,676.2 ± 1,334.1 |
| **Living in the parental home (proportion by age category)** |  |
| Total living in the parental home | 11,387 (89) |
| 10-18 years | 2,374 (99) |
| 19-24 years | 1,889 (84) |
| 25-30 years | 719 (50) |

##### Multilevel models

Data were analysed as multilevel linear regression models with measurement waves nested within individuals. Models 1 and 2 looked at the association between parental FV intake with adolescent FV, adjusting for the covariates outlined above for Model 2. Age categories were added for Model 3 and Model 4 included interaction terms between parental FV intake and age categories. The final model built on Model 4, including the interaction between parental FV intake and whether or not the adolescent was living in the parental home. In order to compare the association between parental FV and adolescent FV at different ages, depending on living arrangement, the dataset was stratified by living arrangement (living in the parental home vs not) and Model 5 analysed in the stratified dataset.

All models were fitted using the *lme4* package in the R statistical environment^55^. Confidence intervals at 95% were computed using the *confint.merMod* method^56^ for multilevel models.

## 4. Results

### 4.1. Sample characteristics and descriptive statistics

The analytical sample consisted of 12,805 adolescents aged 10-30 years that had data on their own FV intake, as well as the FV intake for at least one parent from at least one measurement wave. The mean age of adolescents across all waves was 17.4 years (SD: 5.6) and there was a relatively even split of females and males (52% and 48%, respectively). The majority of the sample was of white ethnicity (72%), which reflects the UK population^57^. Additionally, 52% of parents had at least a degree as their highest educational qualification and net adjusted monthly household income was reported as £1,676.2 (SD: 1,334.1). Almost all adolescents (99%) were living in the parental home at ages 10-18 years, decreasing to 84% at ages 19-24 years and 50% at ages 25-30 years (Table 2).

### 4.2. Change in FV intake

The overall mean adolescent FV intake was 2.92 portions per day (SD: 1.87). When split by age category, mean intake did not differ across different age categories (Appendix 1). Mean parental intake was 3.52 portions per day (SD: 1.95) with little difference seen across 0

### 4.3. Association between parental FV intake and adolescent FV intake

For each portion increase in parental FV per day, adolescent FV intake increased by 0.20 [95% CI: 0.19, 0.22] portions per day, adjusting for sex, ethnicity, parental education, household income and geographic region (see Supplemental File Table S1 for details of all model results).

### 4.4. Changes in the association between parental and adolescent FV intake with age

The association between adolescent FV intake and parental FV intake was lowest at age 10 years (for each portion increase in parental FV intake, there was a 0.12 [95% CI: 0.07, 0.16] portion increase per day of FV), with no change between 10 years and 11-12 and 13-14 years. The association was highest at 17-18 years (β=0.30 [95% CI: 0.27, 0.33]), after which it steadily dropped from ages 21-22 until 27-28 years but remains higher than 10-14 years (Figure 3).

### 4.5. Change in the association with parental FV intake with living arrangement

Compared to those living in the parental home, there was an overall lower association of parental FV intake with adolescent FV intake among those not living in the parental home of -0.10 [95% CI: -0.14, -0.06]. This applies across the different age categories. Figure 4 demonstrates the association of parental FV intake and adolescent FV intake at different age categories, stratified by their living arrangement for adolescents aged 17 years or older (after which more than 1% of adolescents have left the parental home).

## 5. Discussion

### 5.1. Principal findings

This is the first UK-based analysis of the association between parental and adolescent diet across different ages and living arrangements using longitudinal data. We found a positive association between parental and adolescent FV intake at all ages, with the highest association seen at ages 17-18 years and lowest seen in early adolescence (10-14 years). If adolescents do not live in the parental home, the association decreases, however, a change in living arrangement does not overcome the positive association between parental FV and adolescent FV, suggesting a long-term influence of parental behaviours on adolescents even after they have left home. Although the effect size of parental FV intake on adolescent FV intake appears small, at its highest point at age 17-18 years, 0.30 portions per day represents 10% of the average FV intake for adolescents ages 10-30 years, which is equivalent to a 2 portions per week. As previous public health efforts to increase intake in this age group have not been successful, leveraging this positive association between parental and adolescent FV intake when designing interventions and policy may help push adolescents’ intake closer to the recommended intake.

### 5.2 . Strengths and limitations of the study

This analysis makes use of a large panel survey in the UK, which includes both adolescent and parental data. The use of longitudinal data in this analysis allows us to explore the changing association between parents and adolescents at different ages, to understand when parents’ FV intake is most associated with adolescent FV intake. This dataset is unique in that parental data is available at several time points, but has not previously been used to assess associations between diet in adolescents and their parents’ diets and how it changes with age. Although this is not a traditional longitudinal analysis where an exposure at one time point is associated with an outcome at a later timepoint, this analysis based on longitudinal data from the same individuals over time allows incorporation of within-person changes in associations over time, reducing the impact of between-individual confounding.

FV intake is a major component of a healthy diet, and FV intakes have been widely used as an indicator of diet quality^58^, with a considerable body of research showing links between higher FV intakes and a lower risk of adverse health outcomes^11–14,17,23–25^. However the use of FV intake as the main dietary outcome does not provide an overall picture of the diet of the adolescents and is not comprehensive enough to calculate a diet quality score. The analysis could have been strengthened by adding a contrasting ‘unhealthy’ measure of diet or a more comprehensive assessment of diet quality to gain a better understanding of the adolescents’ overall diets, however these measures were not available in this dataset.

However, the measure used in this survey is quick and of low burden to participants. The FV intake measure was self-reported by participants (both adolescents and parents) so there is a chance of bias, for example, social desirability bias which may change depending on age category. The lower association between parental intake and adolescent intake seen in earlier adolescents (10-14 years) could be related to misreporting in younger ages because of a difference in understanding of portion sizes, for example. Collection and analysis of data using more comprehensive measures of diet would be helpful to confirm our findings.

According to the Food Standards Agency, the Covid-19 pandemic changed the UK population’s food habits, including increased time spent on food preparation and eating with family members^59^, which could have contributed to differences in FV intake for adolescents and the association with their parents’ intake seen in our results. The Covid-19 pandemic also changed young people’s living arrangements through their unforeseen return to their parental home^60^. As this analysis includes data that spans through the Covid-19 pandemic, fewer young people may have been living away from the parental home in waves 11-13, which could also explain why there were fewer people living away from the parental home in our analytical sample. However, the accelerated cohort design used in this analysis allows for inclusion of all ages at each wave and therefore, the effects of the pandemic are less likely to have influenced the findings related to change in association between parental FV and adolescent FV intake at different ages.

Due to the sample included in this analysis, it was not possible to incorporate the weights provided with the dataset and therefore, we assume that our analytical sample have equal probabilities of selection and response^61^. However, we are not providing a descriptive analysis of the population but rather looking at associations between exposure and outcome, and we have adjusted for ethnicity and geographic region among our confounder set. The remaining sampling parameters we do not believe to be associated with our exposure and outcome. Additionally, the association between parental FV and adolescent FV may have been driven by other common causes, such as household income, region of residence, and socioeconomic position, as demonstrated in the DAG in Figure 2. These were adjusted for in the covariates included in the models to try to eliminate as far as possible the confounding effects, however as always in observational studies, there remains the possibility of residual confounding.

### 5.3. Comparison with previous evidence and implications of the findings

Existing published longitudinal analyses on the effect of parental behaviours on adolescent behaviours have shown consistency with our finding that there is a positive association between parental and adolescent dietary intake. Lau et al. and Sylvestre et al. both showed a positive association with parental and adolescent FV intakes^42,62^. Two further longitudinal studies in China^41^ and the Netherlands^38^ reinforced the findings of an overall positive association between parental and adolescent dietary intake over time. However, these covered only adolescents up to age 18 years and did not examine the change when they enter early adulthood.

The current study suggests that the association between parental intake and adolescent intake is lower in earlier adolescence, when other factors may take precedence (e.g. peer influence, school meals) and during the transition into early adulthood, parental modelling regains influence on their diets^63^. A similar trend was seen by Arcan et al., showing that parental FV intake predicted FV intakes in young adults (17.1 ± 0.5 years) but not for younger adolescents (12.6 ± 0.6 years) in a US population^40^. Furthermore, psychology studies suggest that time spent with peers increases in early and middle adolescence, peaking at age 14 years^64^ and adolescents aged 13-17 years care more about what their peers think, therefore, are more influenced by them during this time^65^. This is consistent with our findings that the association between parental FV intake and adolescent FV intake is lower from ages 10-16 years. Furthermore, in Knoll et al.’s exploration of social influence on risk taking behaviours in adolescents and young adults showed that younger adolescents (aged 12-14 years) were most influenced by other adolescents potentially due to social conformity, whereas other age groups were more influenced by adults^66^. Although these were different behaviours, they could explain how influences change between younger adolescents and older adolescents, and they reflect our findings of the lower association of parental intake in younger adolescents (10-14 years).

Our findings show that there is a decrease in the association between parental FV intake and adolescent FV intake when they leave the parental home. However, when this is combined with the increase in the association between adolescent FV intake and parental FV intake in the older age categories (17+ years), this results in a continued positive influence of parents on adolescents up to age 30 years. In this sample, 60% of the adolescents are not living in the parental home at age 29-30 years. However, based on the results from Model 5b, the association between parental FV intake and adolescent FV intake was 0.20 [95% CI: 0.14, 0.26] at age 29-30 years, when they are no longer living in the parental home. These results demonstrate that although there is a decline in parental influence after the adolescent has left the parental home, they have a prolonged influence on their intake, regardless of living arrangement. This is reflected in findings from Dickens et al., which showed an association between parental and adolescent intake of healthy and unhealthy foods and that parental modelling of unhealthy snacking behaviour withstood their child moving out to live independently^67^. This enduring association could then continue to influence future generations, when the adolescents have their own children and have a similar influence on their dietary intake through their life.

Overall, this study provides a clearer picture of the trajectory of parental influence with regards to diet in adolescents and young adults, which could inform peak times to intervene with family-centred interventions in older adolescents (17-18 years) living in the parental home or peer-focussed approaches in younger adolescents (10-14 years), keeping in mind that regardless of age, parental intake is positively associated with their child’s intake.

Further studies using longitudinal data on the changing influences of peers in parallel to parents would be required to underpin the findings on changing influences over time seen in this analysis.

## 6. Conclusion

Adolescent FV intake is associated with parents’ FV intake and this relationship changes with age, with the lowest association seen during early adolescence between 10-14 years. When adolescents do not live in the parental home, there is a decline in the association between parental FV intake on their own intake. However, parental FV intake continues to be associated with adolescent FV intake when they are in their 20s and up to 30 years, suggesting that this persists into early adulthood. Therefore, interventions targeting older adolescents could be more effective by involving their parents and when they are still living in the parental home. The influence of positive behaviours could be carried into adulthood and help maintain a higher level of intake that may, in turn, positively influence their own children.

## Data Availability

The data that support the findings of this study are available on request from the UK Data Service, study number (SN) 6614. The data are publicly available, however, they are considered safeguarded and therefore require users to register and accept the End User Licence.

https://doi.org/10.5255/UKDA-SN-6614-19

## Acknowledgements

We would like to acknowledge the contributions of Stephen Sharp for guidance on statistical methods for this analysis, as well as thank the study participants for their contribution in providing the data used in this analysis.

Understanding Society is an initiative funded by the Economic and Social Research Council and various Government Departments, with scientific leadership by the Institute for Social and Economic Research, University of Essex, and survey delivery by NatCen Social Research and Kantar Public. The research data are distributed by the UK Data Service. Fieldwork for the web survey was carried out by Ipsos MORI and for the telephone survey by Kantar.

## 7. Author Contribution Statement

TB conducted the analysis and drafted the manuscript. EMW and JA supported the conceptualization of the study, interpreting results and drafting, reviewing and editing the manuscript. EMW provided additional guidance on the methodology and analysis. All authors reviewed drafts of the manuscript.

---

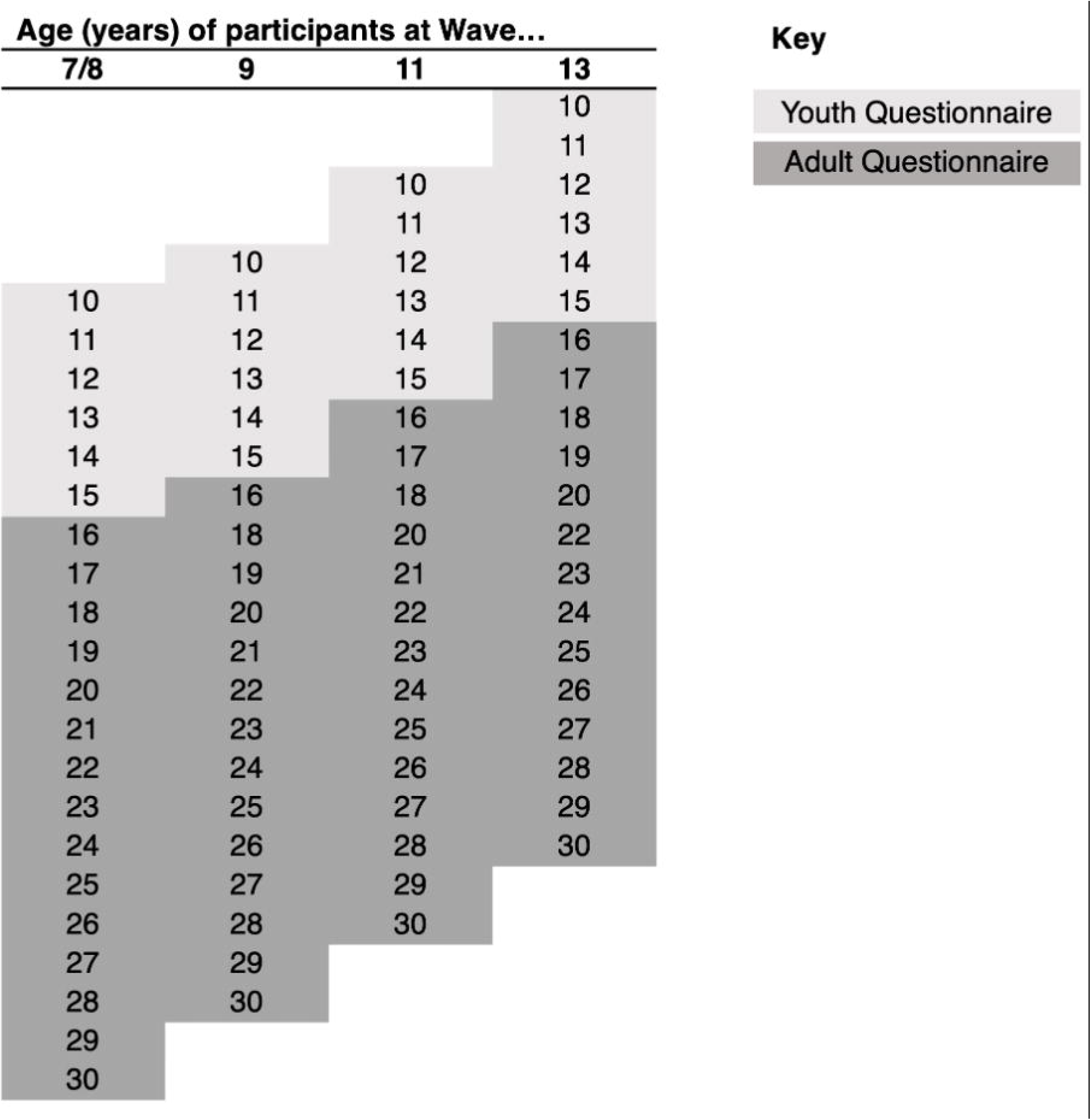

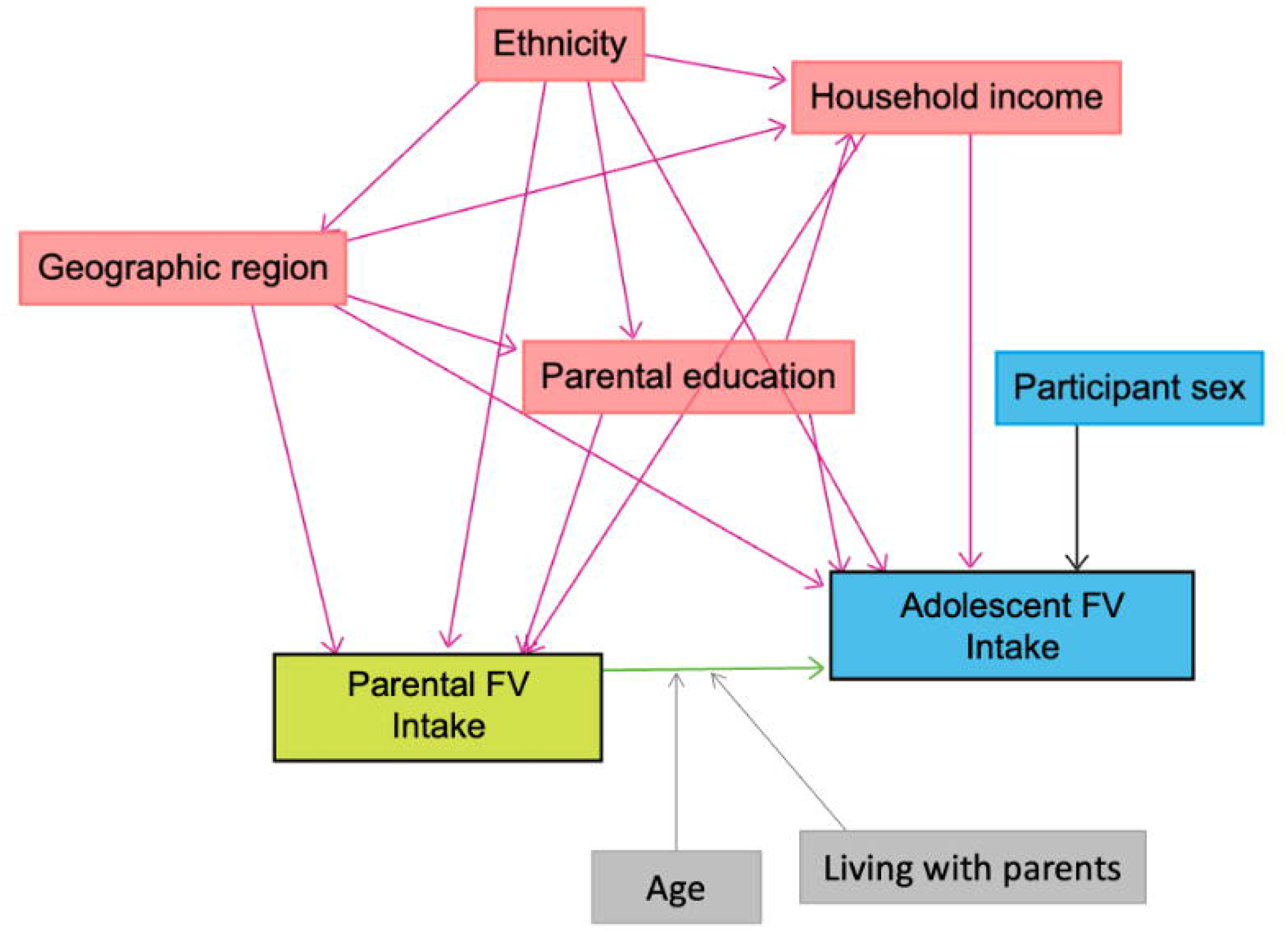

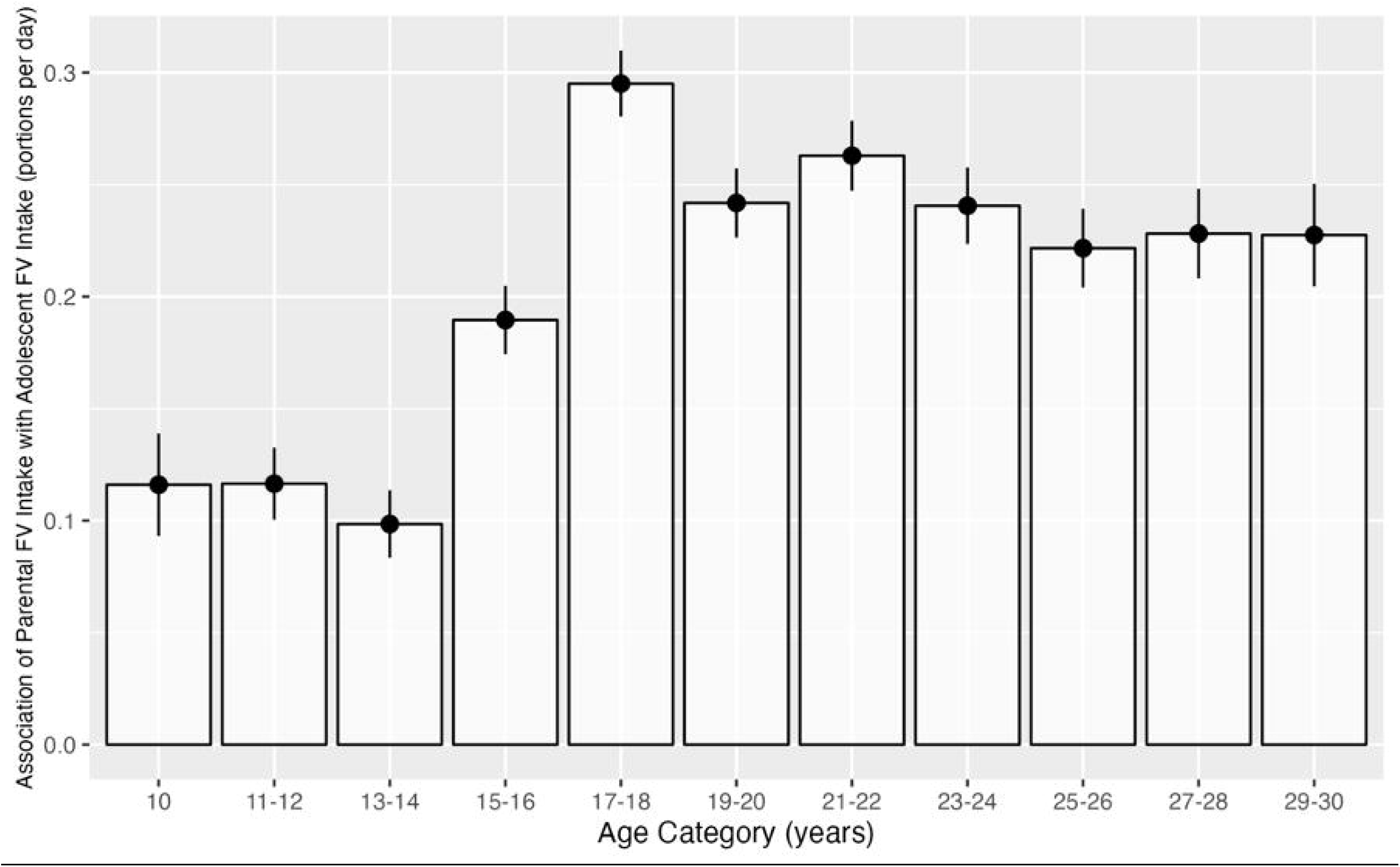

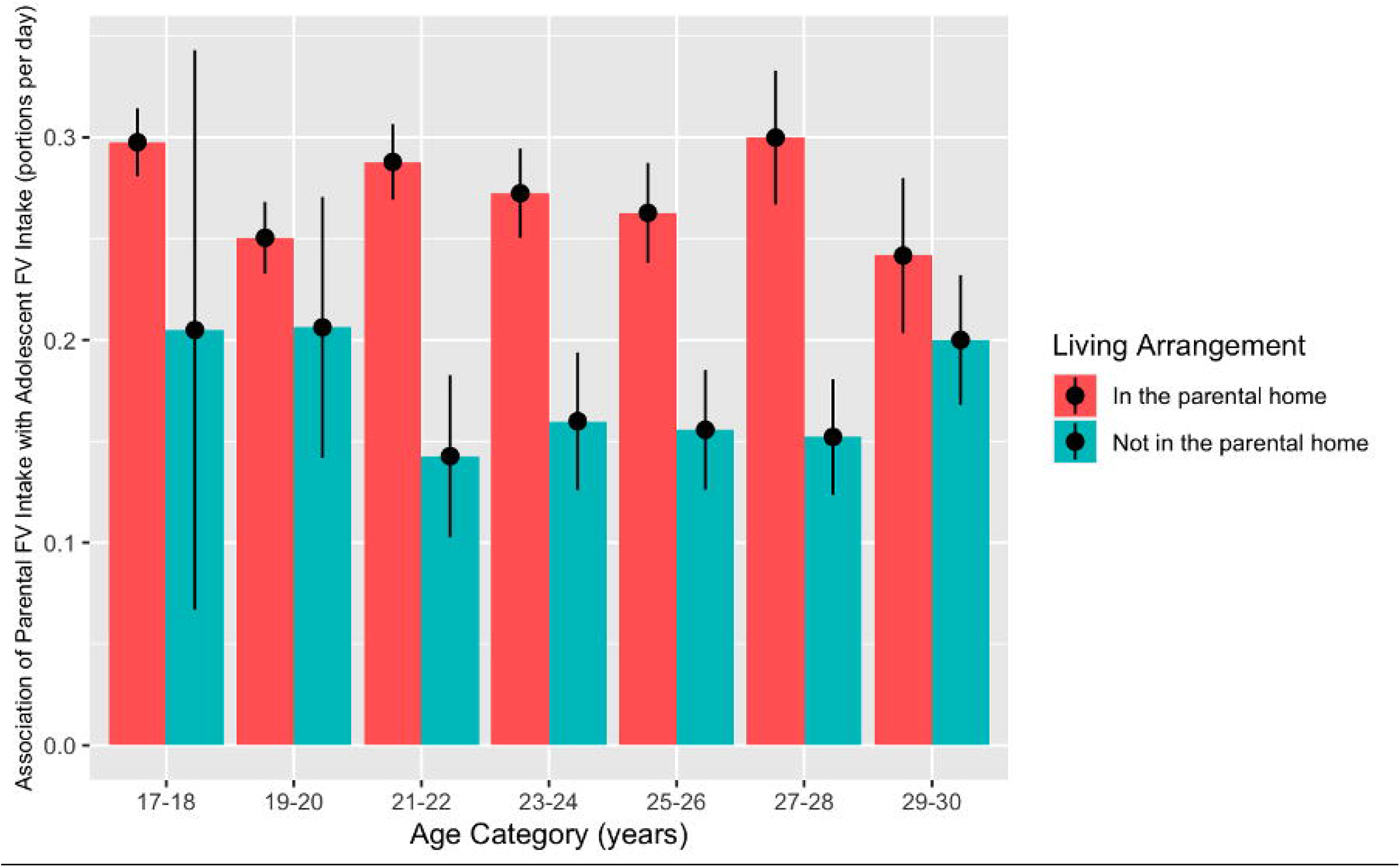

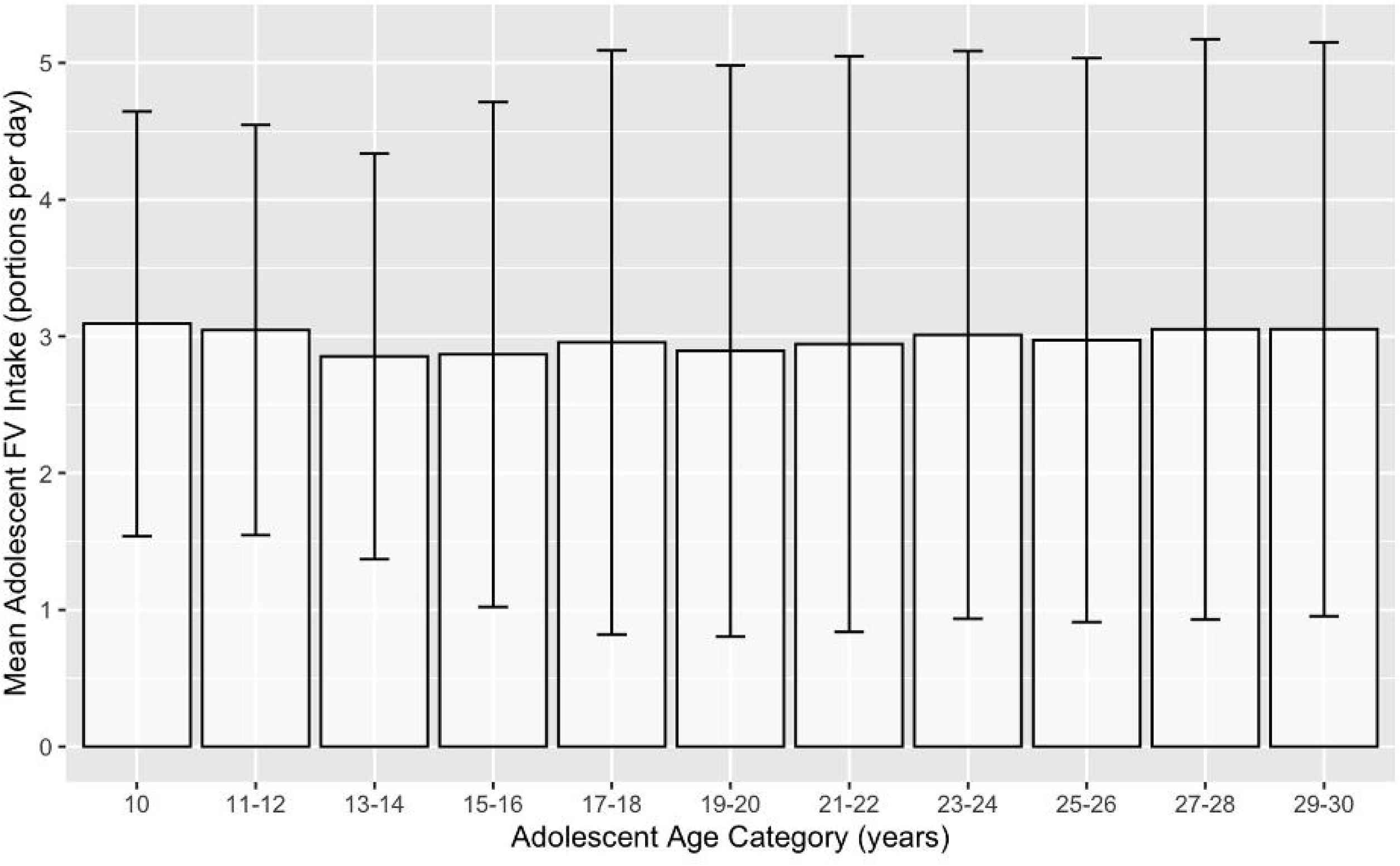

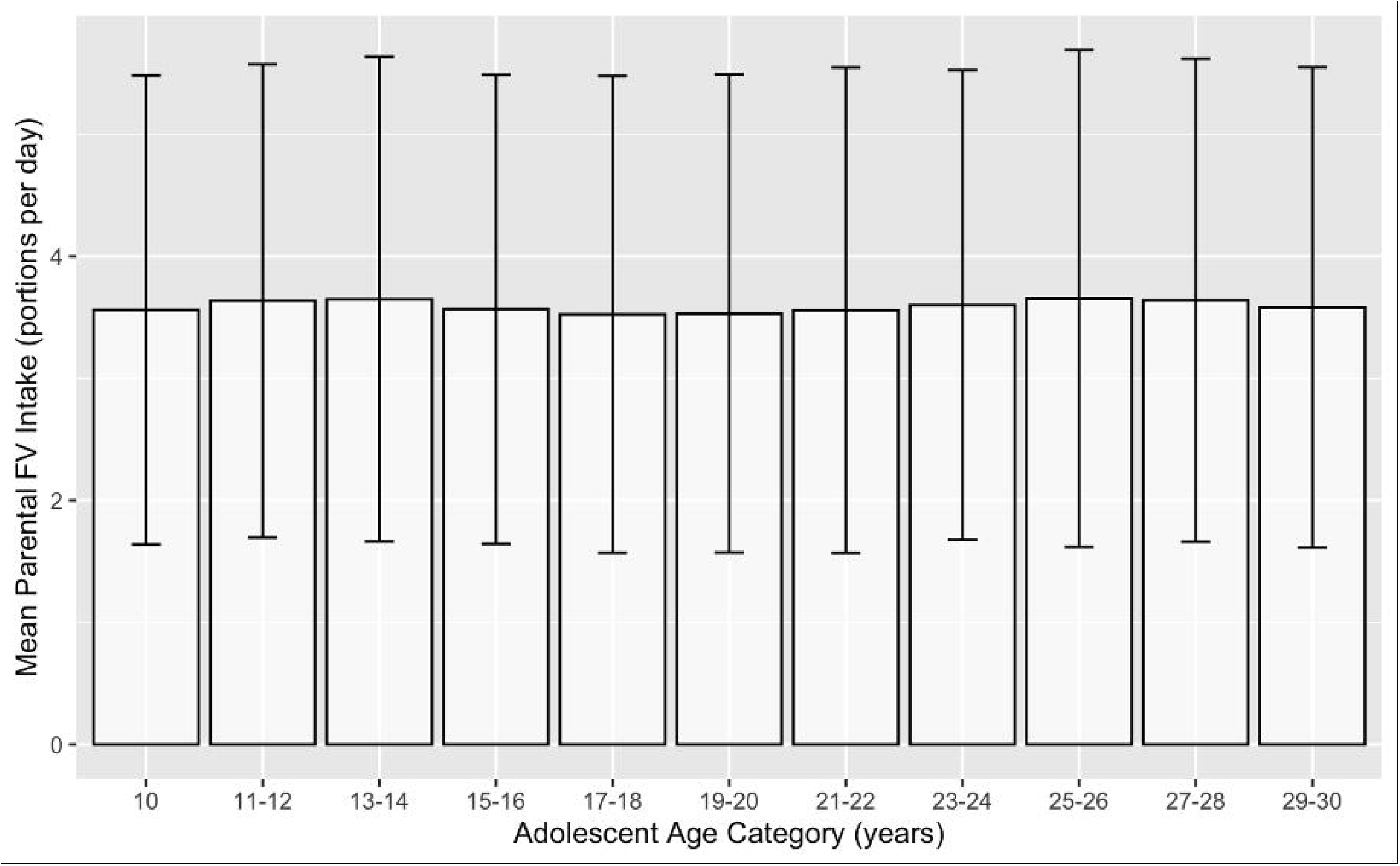

