## Supplemental File Table S1 for "Exploring the changing association between parental and adolescent fruit and vegetable intakes, from age 10 to 30 years"

| | Unadjusted | Adjusted for covariates | Adjusted for age categories | Interactions with age categories | Interactions with living arrangement | Stratified by living arrangement | |
|---|---|---|---|---|---|---|---|
| | **Model 1** | **Model 2** | **Model 3** | **Model 4** | **Model 5** | **Model 5a (living in the parental home)** | **Model 5b (not living in the parental home)** |
| **Parameters** | | | | Estimates [95% CI] | | | |
| (Intercept) | 2.14 *** [2.09, 2.19] | 2.47 *** [2.36, 2.58] | 2.87 *** [2.70, 3.04] | 3.18 *** [2.95, 3.42] | 3.18 *** [2.95, 3.41] | 2.12 *** [1.92, 2.32] | 2.78 *** [1.61, 3.95] |
| Parental FV Intake (portions per day) | 0.23 *** [0.21, 0.24] | 0.20 *** [0.19, 0.22] | 0.20 *** [0.19, 0.22] | 0.12 *** [0.07, 0.16] | 0.12 *** [0.07, 0.16] | 0.30 *** [0.26, 0.33] | 0.20 [-0.07, 0.48] |
| **Covariates** | | | | | | | |
| Sex – Female | | 0.27 *** [0.22, 0.33] | 0.27 *** [0.22, 0.33] | 0.27 *** [0.22, 0.33] | 0.27 *** [0.21, 0.32] | 0.34 *** [0.26, 0.43] | 0.24 ** [0.08, 0.40] |
| Ethnicity – Non-white | | -0.27 *** [-0.35, -0.20] | -0.27 *** [-0.35, -0.20] | -0.28 *** [-0.35, -0.21] | -0.26 *** [-0.33, -0.19] | -0.36 *** [-0.47, -0.26] | -0.43 ** [-0.69, -0.17] |
| Parental education – No degree | | -0.43 *** [-0.49, -0.38] | -0.43 *** [-0.48, -0.37] | -0.43 *** [-0.49, -0.37] | -0.43 *** [-0.48, -0.37] | -0.52 *** [-0.60, -0.43] | -0.50 *** [-0.65, -0.34] |
| Household income (£GBP per month) | | 0.00 *** [0.00, 0.00] | 0.00 *** [0.00, 0.00] | 0.00 *** [0.00, 0.00] | 0.00 *** [0.00, 0.00] | 0.00 ** [0.00, 0.00] | 0.00 * [0.00, 0.00] |
| Geographic Region (ref: London) | | | | | | | |
| North East | | -0.34 *** [-0.51, -0.17] | -0.34 *** [-0.51, -0.17] | -0.35 *** [-0.52, -0.18] | -0.35 *** [-0.51, -0.18] | -0.34 * [-0.60, -0.07] | -0.77 *** [-1.21, -0.33] |
| North West | | -0.23 *** [-0.34, -0.12] | -0.23 *** [-0.34, -0.12] | -0.23 *** [-0.34, -0.12] | -0.23 *** [-0.35, -0.12] | -0.24 ** [-0.41, -0.06] | -0.58 *** [-0.91, -0.25] |
| Yorkshire and the Humber | | -0.26 *** [-0.38, -0.15] | -0.27 *** [-0.38, -0.15] | -0.26 *** [-0.38, -0.15] | -0.27 *** [-0.38, -0.15] | -0.29 ** [-0.47, -0.11] | -0.53 ** [-0.85, -0.20] |
| East Midlands | | -0.12 [-0.25, 0.01] | -0.12 [-0.25, 0.01] | -0.12 [-0.25, 0.01] | -0.12 [-0.25, 0.00] | -0.09 [-0.29, 0.10] | -0.46 ** [-0.80, -0.11] |
| West Midlands | | -0.10 [-0.21, 0.02] | -0.10 [-0.21, 0.01] | -0.09 [-0.20, 0.02] | -0.09 [-0.20, 0.02] | -0.07 [-0.24, 0.10] | -0.40 * [-0.75, -0.05] |
| East of England | | -0.08 [-0.20, 0.05] | -0.08 [-0.20, 0.04] | -0.07 [-0.20, 0.05] | -0.08 [-0.20, 0.04] | 0.03 [-0.16, 0.22] | -0.49 ** [-0.85, -0.14] |
| South East | | 0.01 [-0.11, 0.12] | 0.00 [-0.11, 0.11] | 0.01 [-0.10, 0.12] | 0.01 [-0.10, 0.12] | 0.04 [-0.13, 0.22] | -0.42 ** [-0.73, -0.11] |
| South West | | 0.01 | 0.01 | 0.02 | 0.02 | 0.01 | -0.43 * |

| | | | | | | |
|---|---|---|---|---|---|---|
| | [-0.11, 0.14] | [-0.12, 0.14] | [-0.11, 0.15] | [-0.11, 0.15] | [-0.20, 0.22] | [-0.76, -0.09] |
| Wales | -0.22 ** | -0.22 ** | -0.22 ** | -0.22 ** | -0.14 | -0.50 ** |
| | [-0.36, -0.08] | [-0.36, -0.08] | [-0.36, -0.08] | [-0.36, -0.08] | [-0.36, 0.07] | [-0.86, -0.14] |
| Scotland | -0.16 * | -0.16 * | -0.16 * | -0.16 * | -0.24 * | -0.39 * |
| | [-0.29, -0.03] | [-0.29, -0.03] | [-0.29, -0.03] | [-0.29, -0.03] | [-0.44, -0.03] | [-0.72, -0.05] |
| Northern Ireland | -0.25 *** | -0.25 *** | -0.24 *** | -0.24 *** | -0.29 ** | -0.74 *** |
| | [-0.38, -0.11] | [-0.38, -0.11] | [-0.38, -0.11] | [-0.37, -0.10] | [-0.49, -0.09] | [-1.14, -0.34] |
| **Age category (ref: 10 years)** | | | | | | |
| 11-12 years | | -0.04 | -0.05 | -0.05 | | |
| | | [-0.14, 0.05] | [-0.26, 0.16] | [-0.26, 0.16] | | |
| 13-14 years | | -0.24 *** | -0.17 | -0.17 | | |
| | | [-0.34, -0.14] | [-0.39, 0.04] | [-0.39, 0.04] | | |
| 15-16 year | | -0.36 *** | -0.62 *** | -0.62 *** | | |
| | | [-0.47, -0.24] | [-0.85, -0.40] | [-0.85, -0.40] | | |
| 17-18 years | | -0.42 *** | -1.06 *** | -1.07 *** | | |
| | | [-0.57, -0.27] | [-1.30, -0.82] | [-1.31, -0.83] | **Ref: 17-18 years** | |
| 19-20 years | | -0.48 *** | -0.93 *** | -0.95 *** | 0.10 | 0.11 |
| | | [-0.63, -0.33] | [-1.17, -0.68] | [-1.20, -0.71] | [-0.08, 0.28] | [-1.12, 1.34] |
| 21-22 years | | -0.43 *** | -0.95 *** | -1.02 *** | -0.02 | 0.44 |
| | | [-0.58, -0.29] | [-1.20, -0.71] | [-1.27, -0.78] | [-0.21, 0.17] | [-0.73, 1.61] |
| 23-24 years | | -0.37 *** | -0.82 *** | -0.94 *** | 0.11 | 0.38 |
| | | [-0.52, -0.22] | [-1.07, -0.57] | [-1.20, -0.69] | [-0.10, 0.33] | [-0.78, 1.54] |
| 25-26 years | | -0.43 *** | -0.80 *** | -0.97 *** | 0.10 | 0.29 |
| | | [-0.58, -0.27] | [-1.06, -0.55] | [-1.23, -0.71] | [-0.13, 0.34] | [-0.87, 1.45] |
| 27-28 years | | -0.39 *** | -0.78 *** | -1.01 *** | -0.01 | 0.32 |
| | | [-0.55, -0.23] | [-1.05, -0.51] | [-1.29, -0.74] | [-0.30, 0.28] | [-0.84, 1.48] |
| 29-30 years | | -0.36 *** | -0.78 *** | -1.03 *** | 0.24 | 0.14 |
| | | [-0.53, -0.20] | [-1.06, -0.50] | [-1.32, -0.74] | [-0.09, 0.56] | [-1.03, 1.30] |
| **Interactions (ref: 10 years)** | | | | | | |
| Parental FV X 11-12 years | | | 0.00 | 0.00 | | |
| | | | [-0.05, 0.06] | [-0.05, 0.06] | | |
| Parental FV X 13-14 years | | | -0.02 | -0.02 | | |
| | | | [-0.07, 0.04] | [-0.07, 0.04] | | |
| Parental FV X 15-16 years | | | 0.07 ** | 0.07 ** | | |
| | | | [0.02, 0.13] | [0.02, 0.13] | | |
| Parental FV X 17-18 years | | | 0.18 *** | 0.18 *** | | |
| | | | [0.13, 0.23] | [0.13, 0.23] | **Ref: 17-18 years** | |

| | | | | | | | |
|---|---|---|---|---|---|---|---|
| Parental FV X 19-20 years | | | | 0.13 *** | 0.13 *** | -0.05 * | 0.00 |
| | | | | [0.07, 0.18] | [0.08, 0.19] | [-0.09, -0.00] | [-0.29, 0.29] |
| Parental FV X 21-22 years | | | | 0.15 *** | 0.16 *** | -0.01 | -0.06 |
| | | | | [0.09, 0.20] | [0.11, 0.22] | [-0.06, 0.04] | [-0.34, 0.22] |
| Parental FV X 23-24 years | | | | 0.13 *** | 0.15 *** | -0.03 | -0.05 |
| | | | | [0.07, 0.18] | [0.10, 0.21] | [-0.08, 0.03] | [-0.32, 0.23] |
| Parental FV X 25-26 years | | | | 0.10 *** | 0.14 *** | -0.03 | -0.05 |
| | | | | [0.05, 0.16] | [0.08, 0.20] | [-0.09, 0.02] | [-0.33, 0.23] |
| Parental FV X 27-28 years | | | | 0.11 *** | 0.16 *** | 0.00 | -0.05 |
| | | | | [0.05, 0.17] | [0.10, 0.22] | [-0.07, 0.07] | [-0.33, 0.22] |
| Parental FV X 29-30 years | | | | 0.12 *** | 0.17 *** | -0.06 | -0.00 |
| | | | | [0.05, 0.18] | [0.11, 0.24] | [-0.14, 0.03] | [-0.28, 0.27] |
| Not living with parents | | | | | 0.46 *** | | |
| | | | | | [0.31, 0.61] | | |
| Parental FV X Not living in the parental home | | | | | -0.10 *** | | |
| | | | | | [-0.13, -0.06] | | |
| N | 26687 | 26687 | 26687 | 26687 | 26687 | 12158 | 3958 |
| N (pidp) | 12805 | 12805 | 12805 | 12805 | 12805 | 6979 | 2297 |
| AIC | 104366.28 | 103870.08 | 103819.32 | 103695.90 | 103662.60 | 49270.47 | 16066.14 |
| BIC | 104399.05 | 104033.92 | 104065.08 | 104023.57 | 104006.66 | 49500.05 | 16260.93 |
| R2 (fixed) | 0.05 | 0.09 | 0.09 | 0.09 | 0.09 | 0.12 | 0.07 |
| R2 (total) | 0.49 | 0.50 | 0.50 | 0.50 | 0.50 | 0.53 | 0.60 |

*** $p < 0.001$; ** $p < 0.01$; * $p < 0.05$.

**Table S1:** Multilevel linear regression model estimates. 95% significance level.
